# A Post-Discharge Remote Monitoring System to Enhance Adverse Event Surveillance in Patients with Multiple Chronic Conditions: Design and Field Testing

**DOI:** 10.64898/2026.08.11.26360182

**Authors:** Madeline Smith, Kaitlyn Konieczny, Marie Leeson, Jorge A Rodriguez, Robert S Rudin, Pamela Garabedian, Savanna Plombon, Maria Edelen, Anuj K Dalal

## Abstract

**Background:** Adverse events (AEs) after hospitalization are common and disproportionately affect adults with multiple chronic conditions (MCC). Capturing patient-reported symptoms and self-assessed health may enable earlier detection of post-discharge AEs.

**Objective:** To identify and test user requirements for an automated remote monitoring system to enhance AE surveillance during the transition home following discharge.

**Methods:** We conducted a mixed-methods study using an iterative, user-centered design approach. Semi-structured interviews with patients and clinicians informed system requirements, followed by real-world field testing in 20 patients who used the system for up to 7 days after discharge. The prototype leveraged interoperable electronic health record data services, delivered automated post-discharge check-ins using a combined questionnaire assessing new or worsening symptoms and patient-reported outcomes (PROs), provided risk-stratified health advice (when and with whom to initiate contact), and escalated high-risk symptoms to clinicians in real-time. Descriptive statistics assessed feasibility and utilization; conventional content analysis identified user needs and implementation considerations.

**Results:** Thirty-seven patients with MCC and 23 clinicians participated. Key requirements for patients included clear communication of personalized risk based on red-flag symptoms, and actionable guidance aligned with discharge instructions. Key requirements for clinicians included explicit delineation of responsibility across inpatient and outpatient setting, and selective escalation to minimize burden. Field testing patients completed 60% of the combined questionnaires. Seven patients received Level 2 or Level 3 health advice after reporting new or worsening symptoms. Three patients triggered Level 3 alerts, resulting in one-time, secure escalation emails to clinicians. Four of the 7 patients who received Level 2 or 3 health advice had chart-confirmed emergency department visits within 1 week of discharge. Patients found the system understandable and helpful, while clinicians noted challenges interpreting PRO trends.

**Conclusions:** These observations support the feasibility and acceptability among patients and clinicians of collecting patient-reported symptoms and PROs during the early post-discharge period. Future iterations should prioritize clear risk communication, role clarity, and interpretable patient-reported data. Formal validation is required to assess predictive performance and clinical utility of symptom-based escalation for post-discharge AE surveillance.

## INTRODUCTION

Adverse events (AEs) following hospital discharge pose a significant challenge in healthcare, particularly among populations with multiple chronic conditions (MCC).[1 2] Between 19% and 28% of patients experience AEs after leaving the hospital, leading to increased healthcare burdens through urgent care visits, emergency department admissions, and readmissions.[3–9] These events cause considerable distress and underscore the need for effective care transition interventions in this population. Previous interventions have focused on preventing readmission by improving discharge preparedness, facilitating medication reconciliation, providing adequate home care services and equipment, identifying caregiver and social supports, and ensuring early follow-up with primary care providers.[10–13] Implementing such multifaceted interventions require substantial investment from hospitals and ambulatory practices, with varying impacts on post-discharge outcomes.[13]

Research has shown that hospitalized patients who completed a discharge preparation checklist and subsequently experienced an AE often reported concerning symptoms—such as dyspnea, fever, or chest pain—highlighting the potential for real-time monitoring to detect AEs earlier.[14] Another study indicated that serial administration of global health patient-reported outcomes (PROs) may predict early healthcare utilization events.[15] Collectively, these findings indicate a role for incorporating patient-reported data into AE surveillance. Few studies have examined patient-facing interventions that facilitate routine monitoring of new or worsening symptoms and enable patients to self-assess their general health status during the critical transition period after discharge.[12 14 15]

While various disease-specific remote monitoring systems have emerged for tracking patients’ health status, most have been designed for use in ambulatory settings or target specific conditions or diseases (e.g., asthma, COVID-19), post-surgical populations (e.g., knee replacement, oncologic surgery), or pediatric populations.[16–22] For example, oncology recovery programs offer tools such as questionnaires, historical symptom tracking, and self-management resources, and integrate electronic PROs (ePROs) into electronic health records (EHR) to enhance symptom control and quality of life for cancer patients.[23–26] Scalable remote monitoring systems for asthma or rheumatoid arthritis use ePROs, escalating deteriorating trends to ambulatory providers for triage within EHR workflows.[19 27–29] Few studies have investigated remote monitoring platforms that support patients with MCC and their clinicians after discharge.[30].

## OBJECTIVES

In this study, we used a user-centered design (UCD) approach to identify requirements for an automated post-discharge remote monitoring system for patients with MCC using patient-reported symptoms and self-assessed general health. Based on these requirements, we designed and field-tested a prototype in a real-world setting to evaluate feasibility and acceptability for AE surveillance.

## METHODS

### Study Design, Setting & Participants

We conducted a mixed methods study to identify and evaluate core requirements for the remote monitoring intervention. The study was conducted at Brigham Health affiliated with Mass General Brigham (MGB), a large academic medical center in Boston, MA, which uses a vendor EHR (Epic Systems, Inc.). Eligible patients were adults aged 18 years or older with two or more chronic conditions (hypertension, diabetes mellitus, chronic kidney disease, etc.) who were hospitalized on a general medicine service for more than 24 hours. Eligible patients needed to be English-speaking or have an English-speaking healthcare proxy who could participate on their behalf. Eligible clinicians included discharging attendings (i.e., the physician responsible for inpatient care at discharge), primary care physicians (PCPs), and advanced practice providers (APPs). The MGB Institutional Review Board reviewed and approved this study as minimal risk.

### Sampling & Consent

Eligible patients were approached in the hospital by a research assistant (RA) and were enrolled upon providing verbal informed consent for data collection (semi-structured interviews or post-discharge questionnaires). Prior to field testing, hospital attendings and responding clinicians were informed about the study during grand rounds. During field testing, we notified discharging attendings, responding clinicians, and PCPs of their patients’ enrollment and invited them to participate by email. Patients and clinicians were offered a $40 gift card to participate in data collection activities.

### Overview of Remote Monitoring Intervention

The intervention’s overall design was informed by our prior research utilizing the Non-adoption, Abandonment, Scale-up, Spread, and Sustainability (NASSS) framework to assess the complexities associated with implementing remote monitoring in ambulatory care and emerging literature on remote patient monitoring.[17–19 31–33] This approach marked a shift from conventional post-discharge care to a digitally enabled paradigm (**Figure 1**).[19 34] The NASSS framework guided attention to the condition burden of patients with MCC, the complexity of patient- and clinician-facing workflows, the need to align escalation protocols with existing care transitions responsibilities, and organizational constraints associated with integrating remote monitoring into EHR-based communication channels, emphasizing simplicity in the design process.[33]

**Figure 1.**
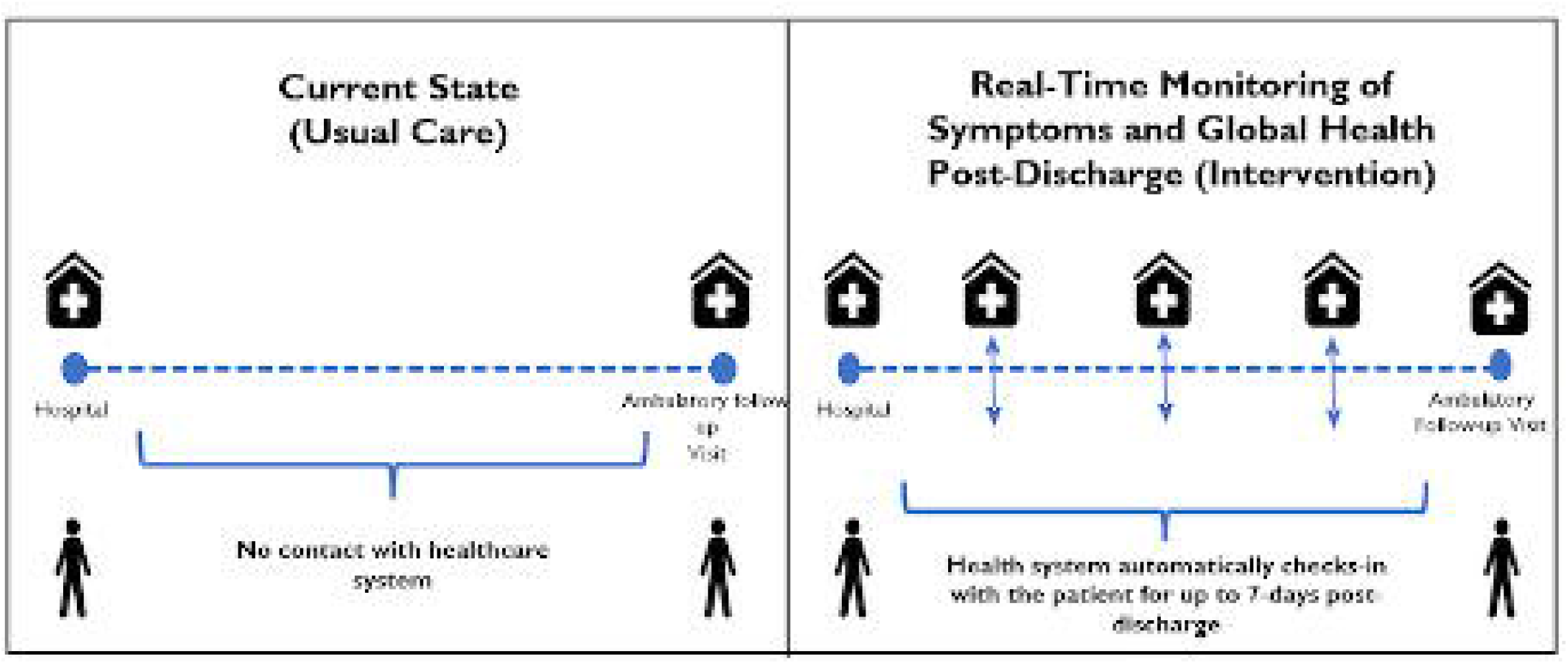
Current and anticipated future state. Transition from traditional post-discharge ambulatory care to a digitally enabled remote monitoring model. Real-time monitoring of new and worsening symptoms and health outcomes (PROMIS Global-10) reported by the patient can be facilitated by automated daily check-ins. Other routine care activities, like post-discharge phone calls being adopted across primary care clinics as part of value-based care initiatives could also be incorporated.[55]

Consistent with our research objectives, two primary areas were addressed: (1) preliminary testing of an escalation protocol for early detection of post-discharge AEs through patient-reported new or worsening symptoms modeled on reports in other hospital populations and prior analytic work conducted at our institution;[14 17 31 32] and (2) the integration of patient-reported outcomes (PROs) to support a thorough evaluation of patients’ health status using the validated PROMIS Global-10 questionnaire, which examines physical, mental, and social dimensions pertinent to the complex needs of our patient population.[15 35 36]

### Used-Centered Design

We conducted semi-structured interviews tailored to patients or clinicians across two phases. Interview questions (**Supplementary Materials**) focused on six distinct pillars of a high-quality care transition: discharge planning, promoting self-management, enlisting social and community supports, coordinating care among team members, monitoring symptoms after discharge, and outpatient follow-up.[37] We anticipated recruiting 30 patients and 20 clinicians across both phases.

#### User Needs & Requirements Gathering

First, we identified user needs and gathered preliminary requirements aligning with our research objectives. For patients approached in the hospital, an RA asked a series of questions related to their expectations about the discharge process and transition home. The RA held a 30-minute follow-up with patients 1-2 weeks after discharge, asking about their hospital discharge experience, recovery, and available support. For clinicians, the RA asked a series of questions about their experience discharging patients, typical workflow before and after discharge, and how they discuss health risks or AEs with patients.

#### Prototype Creation

Next, design solutions were created to meet new user needs, leading to an initial REDCap prototype, which was called MyPostDischargePal (**Figure 2**). The prototype triggered self-management advice based on after-visit summary (AVS) instructions and questionnaire responses. Application programming interfaces (API) that use the Fast Health Interoperability Resources (FHIR) standard were identified to extract EHR data corresponding to health information from the AVS. A combined questionnaire, initially configured to be sent daily for up to 7-days after discharge, included daily symptom checks (Memorial Symptom Assessment Scale) and the PROMIS Global-10 administered on days 1, 3, 5, and 7. The escalation framework—informed by remote monitoring systems reported in literature, and expert advice from our study’s advisory committee—was structured to have three tiers of advice.[17 18 31 32] After automated daily check-in during which patients reported symptoms, a dynamic risk score was computed based on counts of red flag and non-red flag symptoms. Risk scores were pragmatically assigned by expert-informed consensus given the focus on feasibility testing; thresholds were not prospectively validated. Examples of new and worsening symptoms considered red flags included GI issues, shortness of breath, swelling, chest pain, or fever which had been identified as predictors of post-discharge AEs in our prior research.[12 14]

**Figure 2.**
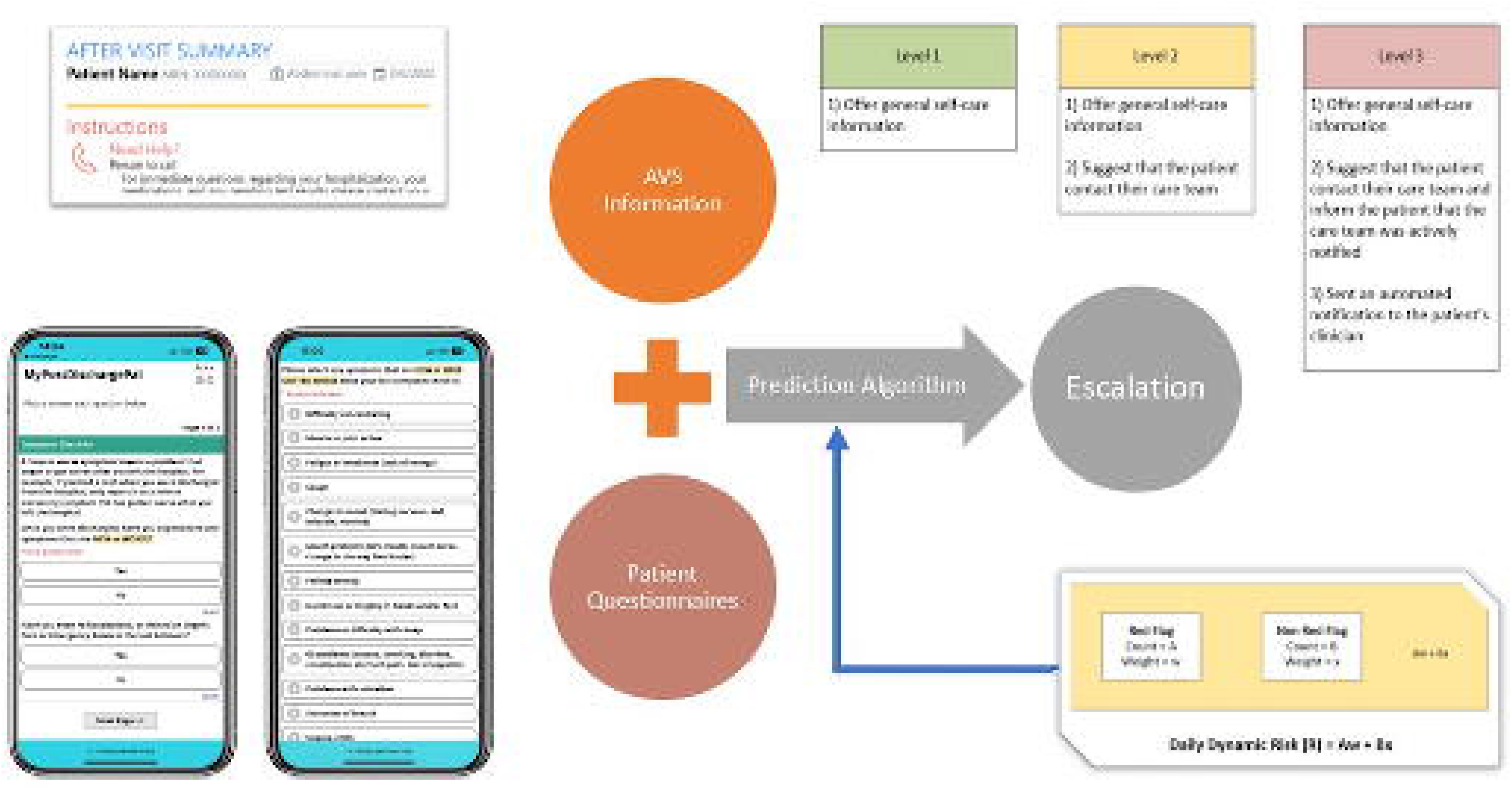
MyPostDischargePal – App Prototype. The app prototype, developed in REDCap, integrated pertinent health data from the AVS and utilized a consolidated questionnaire derived from the Memorial System Assessment Scale and Global-10 PROMIS questionnaires. A prediction algorithm calculated a daily dynamic risk score to trigger three levels of health advice. When a patient reported new or worsening symptoms, tailored health recommendations— aligned with predefined risk thresholds—were sent via email.

#### Iterative Prototype Refinement

The prototype was iteratively refined over ten rounds with the research team to simulate questionnaire flow, evaluate EHR data retrieval reliability, adjust logic, and clarify content of automated emails sent to patients and clinicians. Once core user requirements were satisfactorily addressed, a subsequent round of testing was conducted with patient advisors. The enrollment email, questionnaires, check-in process, and health advice notifications were reviewed during an initial meeting. During a mock role-playing scenario, participants interacted with the prototype for up to seven days following discharge. Feedback was solicited during the initial meeting and upon completion of the scenario via follow-up email or phone call. Modifications to the prototype were made based on the analysis of feedback.

#### Field Testing

We conducted field testing using the REDCap prototype to evaluate the feasibility during the early post-discharge period based on initial user requirements. Patients were contacted and asked to participate in a semi-structured interview upon completion of the 7-day monitoring period. They were asked about their perception of the intervention, including their experience completing questionnaires and reporting symptoms. Clinicians of enrolled pilot patients were asked to participate in a semi-structured interview (discharging attendings and PCPs). In addition, a working group of hospital-based clinicians met five times during this time to provide feedback. Feedback from interviews and working group members was used to further refine the intervention prior to the main trial.

### Data Collection

Demographic data were obtained from patients, patient advisors, and clinicians who participated in interviews, and clinicians participating in the working group. Information from the AVS—including which symptoms to report, when and whom to call, and educational materials— was extracted from the EHR for all patients participating in field testing. Patient questionnaires were administered using REDCap. Administrative EHR data, such as length of stay (LOS) and post-discharge encounters, were collected through chart extraction. All interviews were recorded using a secure audio recording application (Microsoft Teams).

### Analysis

We used descriptive statistics to summarize participant demographics, AVS content, questionnaire completion rates, and change in physical and mental health scores derived from the PROMIS Global-10 questionnaire. Audio recordings were transcribed and interview transcripts were independently reviewed and coded by two members of the research team in Excel (Microsoft, Inc.). Coders were not blinded to participant type (patient or clinician), as this context was relevant to interpretation of responses. Conventional content analysis was used to identify concepts and group codes into emerging user requirements. Coding was conducted iteratively, with regular meetings between coders to compare interpretations, discuss discrepancies, and refine the coding framework until consensus was reached. Representative quotations were selected to illustrate major themes. We anticipated achieving thematic saturation after 12–15 patient interviews and 10–12 clinician interviews during the needs-gathering phase and after 10–12 patient interviews and 8–10 clinician interviews during field testing. Final themes and user requirements were reviewed and confirmed through team-based consensus and subsequently used to guide prototype design and refinement. Feedback and additional user needs identified during field testing were incorporated into the final intervention.

## RESULTS

A total of 37 patients (**Table 1**) and 23 clinicians participated. Of the 37 patients (10 [27.0%] were aged ≥65, 21 [56.8%] were female, 25 [67.6% ]were White, 32 [86.5%] were non-Hispanic), 17 participated in user needs gathering interviews and 20 participated in field testing. The mean (SD) and median (IQR) number of chronic conditions was 4.2 (2.1) and 4 (3), respectively. The most frequent conditions in descending order included hypertension (45.9%), depression (40.5%), anxiety (29.7%), diabetes (27.0%), asthma (27.0%), arthritis (27.0%), hyperlipidemia (24.3%), cancer (10.8%), osteoporosis (10.8%), chronic kidney disease (10.8%), heart failure (8.1%), end-stage renal disease (5.4%), chronic obstructive pulmonary disease (5.4%), and coronary artery disease (2.7%).

**Table 1.** Patient Characteristics.

| **Characteristic** | **All, n=37\*** | **Needs-gathering, n=17** | **Field Testing, n=20<sup>‡</sup>** |
| --- | --- | --- | --- |
| **Unique participants – no. (%)** | 37 (100) | 17 (100) | 20 (100) |
| **Age – no. (%)** |  |  |  |
| <65 | 27 (73.0) | 13 (76.5) | 15 (75.0) |
| >=65 | 10 (27.0) | 4 (23.5) | 5 (25.0) |
| **Gender – no. (%)** |  |  |  |
| Male | 16 (43.2) | 8 (47.1) | 8 (40.0) |
| Female | 21 (56.8) | 9 (52.9) | 12 (60.0) |
| **English Language Abilities – no. (%)** |  |  |  |
| English speaking | 37 (100) | 17 (100) | 20 (100) |
| Non-English speaking* | 0 (0) | 0 (0) | 0 (0) |
| **Race – no. (%)** |  |  |  |
| White | 25 (67.6) | 12 (70.6) | 13 (65.0) |
| Black or African American | 8 (21.6) | 4 (23.5) | 4 (20.0) |
| Asian | 1 (2.7) | 0 (0) | 1 (5.0) |
| Other | 3 (8.1) | 1 (5.9) | 2 (10.0) |
| **Ethnicity – no. (%)** |  |  |  |
| Hispanic | 5 (13.5) | 3 (17.6) | 2 (10.0) |
| Non-Hispanic | 32 (86.5) | 14 (82.4) | 18 (90.0) |
| **Chronic Conditions – no. (%)** |  |  |  |
| 2 | 7 (18.9) | 4 (23.5) | 3 (15.0) |
| 3 | 7 (18.9) | 2 (11.8) | 5 (25.0) |
| 4-5 | 11 (29.7) | 3 (17.6) | 8 (40.0) |
| 6-10 | 7 (18.9) | 3 (17.6) | 4 (20.0) |
| Missing* | 5 (13.5) | 5 (13.5) | 0 (0.0) |
| **Chronic Conditions – no. (%)** |  |  |  |
| Depression | 15 (40.5) | 5 (29.4) | 10 (50.0) |
| Anxiety | 11 (29.7) | 3 (17.6) | 8 (40.0) |
| Hypertension | 17 (45.9) | 10 (58.8) | 7 (35.0) |
| Diabetes | 10 (27.0) | 4 (23.5) | 6 (30.0) |
| Asthma | 10 (27.0) | 4 (23.5) | 6 (30.0) |
| Hyperlipidemia | 9 (24.3) | 3 (17.6) | 6 (30.0) |
| Arthritis | 10 (27.0) | 4 (23.5) | 6 (30.0) |
| Cancer | 4 (10.8) | 1 (5.9) | 3 (15.0) |
| Osteoporosis | 4 (10.8) | 2 (11.8) | 2 (10.0) |
| Substance use disorder | 4 (10.8) | 2 (11.8) | 2 (10.0) |
| Chronic kidney disease | 2 (5.4) | 1 (5.9) | 1 (5.0) |
| End-stage renal disease | 2 (5.4) | 1 (5.9) | 1 (5.0) |
| Coronary artery disease | 1 (2.7) | 0 (0) | 1 (5.0) |
| Chronic obstructive pulmonary disease | 2 (5.4) | 1 (5.9) | 1 (5.0) |
| Heart failure | 3 (8.1) | 2 (11.8) | 1 (5.0) |
| Missing* | 5 (13.5) | 5 (29.4) | 0 (0) |

Conditions from AHRQ's multiple chronic condition list included: depression, anxiety, hypertension, diabetes, asthma, hyperlipidemia, arthritis, cancer, osteoporosis, substance use disorder, chronic kidney disease, end-stage renal disease, coronary artery disease, chronic obstructive pulmonary disease, heart failure
\*Chronic condition data were unavailable for 5 patient advisors.
<sup>‡</sup>After visit summary data are only reported for the 20 field-testing patients: mean (SD) page length: 12 (4.07); discharging attending designated the 'person to call', 13 (65%); fewer than 5 'when to call' symptoms documented, 11 (65%); educational materials specific to admission or discharge diagnosis provided, 6 (30%)

Of the 23 clinicians, 12 participated in user needs interviews and 11 participated in field testing: 3 (13.0%) were aged ≥65, 17 (73.9%) were female, 13 (56.5%) were White, 13 (56.5%) were physicians, 10 (43.5%) were advanced practice providers, 14 (60.9%) had less than 10 years of experience.

### Requirements Informed by User Needs

Many patients (**Table 2, top**) reported needing clear information about personal health risks, delivered in simple, accessible language tailored to their medical knowledge to understand their recovery needs at discharge. While some felt that digital tools like patient portals could improve transparency and communication of specific actionable steps, many still preferred phone calls for timely responses from doctors.

**Table 2.** User Needs & Requirements from Patients and Clinicians.

| User Needs & Requirements | Quote |
| --- | --- |
| **Patients** |  |
| Health risks should be clear and transparent to better understand and manage their recovery. | *"Yes, [I would be interested in understanding my risk of something going wrong]. You never know what could really happen in the next 30 days, because you could be fine or something drastic could happen."* |
| Information should be explained in simple, accessible language. | *"If they [clinicians] talk about risk factors, it doesn't scare me it just makes me take things more seriously."* |
| Specific actionable steps for high-risk patients for effective self-management. | *"I'm reading the part that says you have a medium risk of something going wrong in the next 30 days. That might scare me a little bit... as long as there is information, like there is here, on what to do based on your risk factor, I think that would be helpful... I would want very specific information about what might happen [and] what I need to do immediately at that moment?"* |
| Digital tools should support the efficient submission of questionnaires and communication with doctors. | *"Make it user-friendly... Like, don't make it so it takes me forever to do you know?... The shorter the better"* |
| **Clinicians** |  |
| Any patient, regardless of device or tech skills, should be able to understand and mitigate their risks. | *"....you have to have a smart phone, and you have to download an app. People who don't have those things and aren't like tech savvy or don't have any family support, like those are the people who are most at risk for adverse effects. And this doesn't reach them, which is always a limitation of our technology-based interventions."* |
| Clear responsibility for post-discharge care to ensure timely support and guidance for patients. | *"I think it's my professional and moral responsibility to be available to them if they are getting worse from something that I managed and said I thought they were OK to go home... If it's not an urgent issue, it should go to their PCP to continue to manage because then it's no longer appropriate for me to continue to meddle so often."* |
| Clear communication protocols with PCPs to improve care coordination and support continuity. | *"...if they're out of the system, it's very hard to get in direct contact. If you wanted to talk to the PCP or any outpatient provider, you're stuck calling their office and you get the nurse, and then they say, oh, we'll relay the message and then maybe in the next day, you'll hear from them... I think if there was like a way to just like electronically communicate, it would be a lot easier, I think without other members of the care team. I mean, just send a quick message."* |
| A smart system to manage symptom notifications without overwhelming clinicians, ensuring patient needs are met efficiently. | *"I think that there are patients that will be using it constantly and I find that that gives me anxiety. Thinking about them Patient Gateway messaging me all the time to be like 'ohh my gosh I just took my blood pressure and it's 100 over' [or] 'what should I do with my medications?'... That gives me a little bit of anxiety thinking about them."* |

Clinicians (**Table 2, bottom**) rarely used the term “adverse event”, instead discussing specific risks tied to their patients’ conditions and treatments. They considered factors like mental and social status, communication skills, education, frailty, age, comorbidities, medications, and follow-up needs. Many assumed the responsibility for discharge planning, including preparing care instructions and, at times, scheduling follow-up. Responsibility for post-discharge care was split: some clinicians favored PCP involvement, while others suggested continued discharging attending accountability for up to 72 hours. Lastly, a ‘smart’ alerting system to address concerns about excessive notifications for non-urgent symptoms and difficulties contacting out-of-network PCPs was favored.

### Prototype Refinements

The data retrieval methods, questionnaire design, content, and flow (**Figure 3**), and escalation logic in the initial configuration of the REDCap prototype were iteratively refined to align with emerging patient and clinician requirements (**Table 2**). For example, to minimize patient burden, questionnaires were kept concise and used straightforward language. Essential EHR data—including discharging attending, PCP, current medications, scheduled follow-up appointments, pending test results, and dietary restrictions—were mapped to FHIR standards (**Supplementary Materials**) and automatically retrieved using a Python script integrated with REDCap’s API. Lastly, patient emails included hyperlinks to enterprise portal messaging, direct phone numbers, and upcoming appointments.

**Figure 3.**
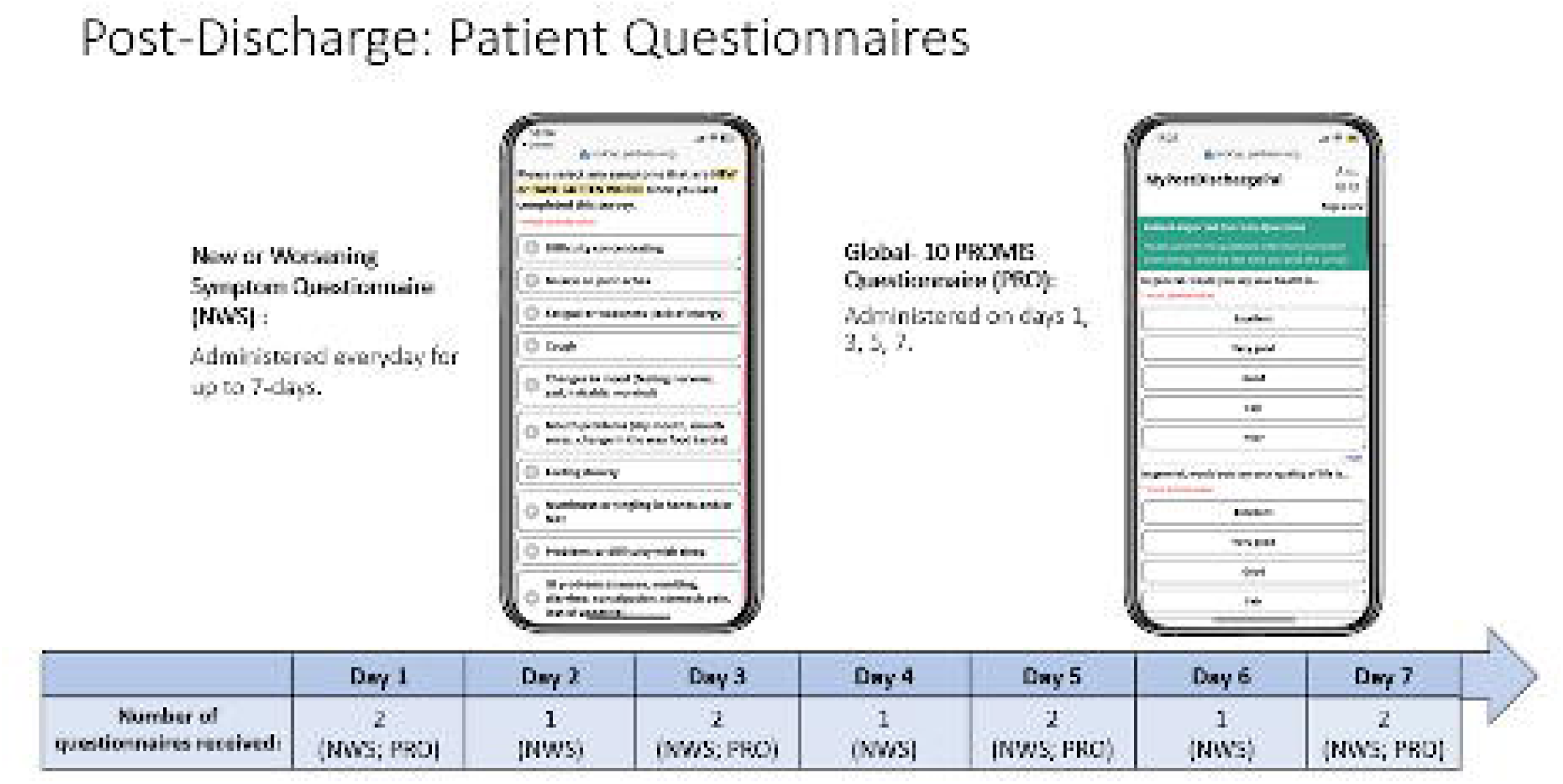
Questionnaire design and flow. The app facilitated patient self-reporting of new or worsening symptoms (NWS) and overall health status using 10-item global health patient-reported outcome (PRO). Symptom questionnaires were administered daily over a seven-day period, while global health assessments were scheduled for Days 1, 3, 5, and 7 as add-ons to symptom questionnaires. On Day 2, patients also received a post-discharge check-in questionnaire aligned with the institution’s phone call protocol, in addition to the symptom questionnaire. In total, each patient could receive up to 7 NWS and 4 PRO questionnaires over the 7-day monitoring period. If a patient indicated they were in the emergency department, future questionnaires were suppressed.

Drawing from reported experience of other investigators, a scoring system was established to guide escalation logic (**Table 3**), where patient-reported symptoms were pragmatically assigned weighted scores of one or two points.[17 31 32] For the target patient population, symptoms (such as gastrointestinal issues, shortness of breath, chest pain, swollen legs, fever, etc.) which were associated with a high likelihood of post-discharge AEs in our prior multivariable analysis were classified as ‘red flags’ and assigned a score of two points each, while all other symptoms received one point.[14] The cumulative symptom score triggered health advice to the patient at three predefined risk thresholds: Level 1 for scores greater than 0 and up to 1, Level 2 for scores greater than 1 and up to 3, and Level 3 for scores greater than 3. For the highest risk tier (Level 3), a notification was also sent to the discharging attending and PCP (if affiliated with MGB) via secure email. Lastly, if a patient indicated an emergency department visit or re-admission, the system automatically halted further questionnaire distribution.

**Table 3.** Health advice levels and risk score thresholds based on new or worsening symptoms.

| **Risk score thresholds** | **Health Advice** |
| --- | --- |
| *Health Advice Level 1: $0 < R \leq 1$* | *If symptoms persist or worsen, contact your doctor* |
| *Health Advice Level 2: $1 < R \leq 3$* | *We recommend contacting your doctor* |
| *Health Advice Level 3: $R > 3$* | *Contact your doctor. Your doctor was notified* |
| **Red flag symptoms: 2 points** |  |
| • Cough<br>• Gastrointestinal (GI) problems<br>• Shortness of breath<br>• Dizziness, lightheadedness, fainting<br>• Swollen arms or legs | • Headache<br>• Falls or trouble with balance<br>• Chest pain<br>• Bleeding<br>• Fever |
| **Non-red flag symptoms: 1 point** |  |
| • Difficulty concentrating<br>• Muscle or joint aches<br>• Fatigue or weakness<br>• Changes in mood<br>• Mouth problems<br>• Feeling drowsy<br>• Numbness or tingling in hands or feet | • Problems or difficulty with sleep<br>• Problems with urination<br>• Sweats/chills<br>• Problems with sexual function<br>• Changes in skin<br>• Lack of appetite or weight loss<br>• Difficulty swallowing |
| *R = Total symptom score |  |

Health advice (**Figure 4**) about patient-reported new or worsening symptoms was communicated at three risk levels (Level 1, 2, 3) via email subject headers with corresponding color-coded banners (green, yellow, red) in the email body aligned with our health networks patient communication standard. Red-flag symptoms appeared bolded: “you reported the following symptoms that were considered high risk: **chest pain**, muscle aches.” The content of the body also included key medication changes, contact details, and follow-up appointments from the AVS. Clinician notifications were sent to the discharging attending and carbon-copied the network PCP to clarify responsibility.[38–40] To limit email burden, escalation emails were sent to clinicians on the first day patient reported symptoms triggering risk level 3 health advice. All notification activity were carbon-copied to the research team’s centralized email inbox to ensure correct wording and appropriate triggering of all health advice to patients and escalation notifications to clinicians as required by our IRB protocol.

**Figure 4.**
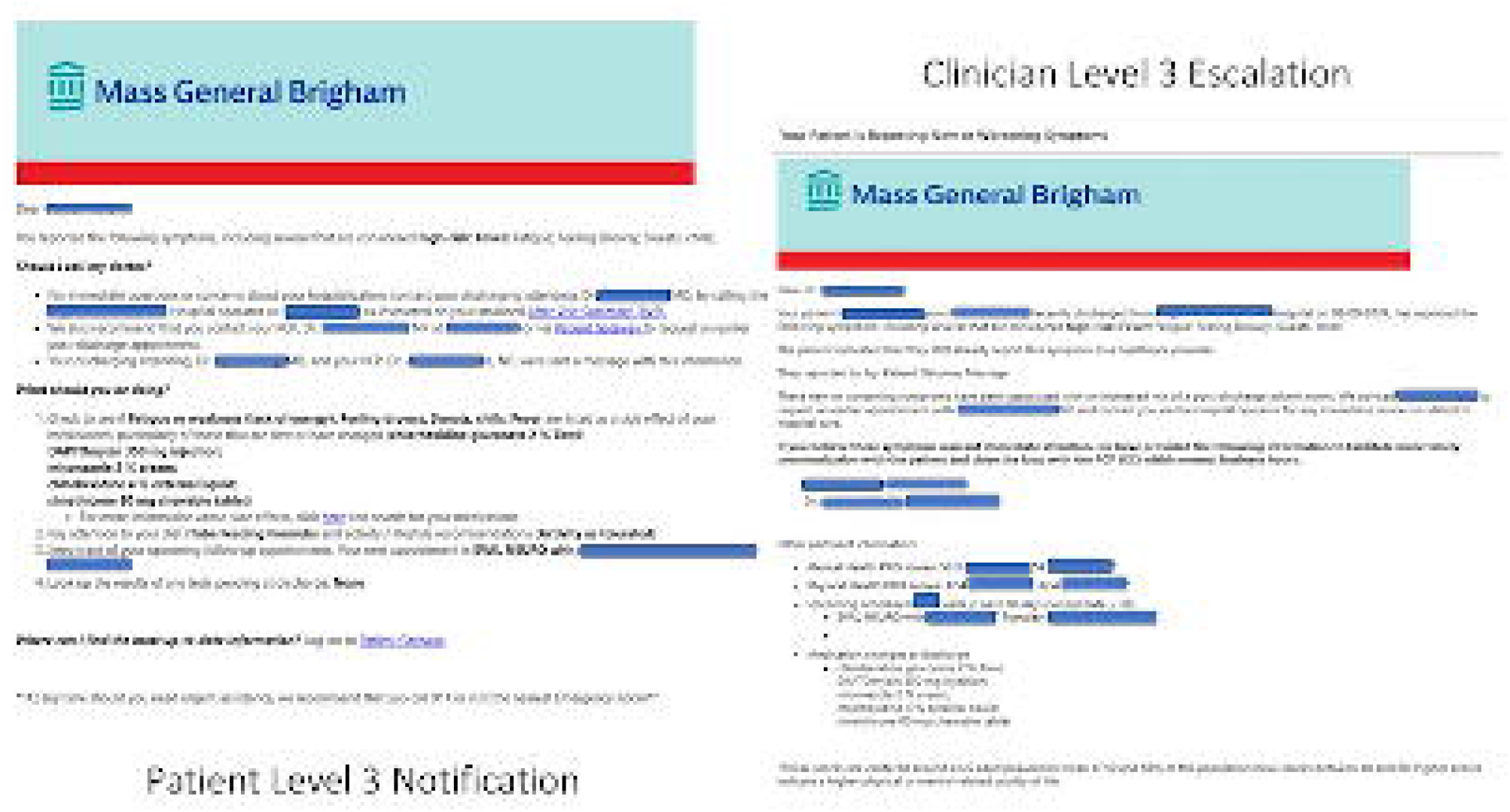
Risk Level 3 health advice received by patient (left) and corresponding Level 3 escalation notification sent to clinicians (right). Email notifications were triggered by the pragmatic risk score calculated based on number and type of patient-reported new or worsening symptoms. The discharging attending (TO) and PCP (CC) were notified when patient-reported symptoms triggered risk level 3 health advice (dynamic risk score > 3). To minimize email burden, escalation emails were sent to clinicians only the first time a patient-reported symptom triggered risk level 3 health advice.

### Field Testing

For the 20 patient participants (**Table 4**), a total of 140 questionnaires were available (140 symptom, 80 PRO) and 130 were sent (130 symptom, 75 PRO) during the 7-day post-discharge monitoring period. Ten questionnaires (10 symptom, 5 PRO) were not sent to four patients who reported being in the emergency room: two on Day 3, one on Day 5, and one on Day 7. Of the 15 patients completing at least one questionnaire, 3 (20%) were aged ≥65, 8 (53.3%) were female, 11 (73.3%) were White, and 13 (86.7%) were non-Hispanic. Of the five patients who did not complete any questionnaires, 1 was aged ≥65 years, 1 was female, 2 were White, and 2 were non-Hispanic.

**Table 4.** Field-testing feasibility and utilization outcomes.

| **Measure, n (%)** | **Result** |
| --- | --- |
| Field-testing participants | 20 |
| Symptom questionnaires available | 140 |
| PRO questionnaires available | 80 |
| Symptom questionnaires sent | 130 |
| PRO questionnaires sent | 75 |
| Symptom questionnaires completed | 78/130 (60.0%) |
| PRO questionnaires completed | 45/75 (60.0%) |
| Patients completing $\geq 1$ questionnaire | 15/20 (75.0%) |
| Patients completing no questionnaires | 5/20 (25.0%) |
| Patients receiving any health advice | 7/20 (35.0%) |
| Level 1 health advice recipients | 3/20 (15.0%) |
| Level 2 health advice recipients | 2/20 (10.0%)* |
| Level 3 health advice recipients | 3/20 (15.0%)* |
| Level 3 escalation emails sent | 3 |
| ED visits within 7 days among Level 2/3 recipients | 4/7 (57.1%) |
| ED visits within 7 days among patients without symptoms/no submission | 0/13 (0.0%) |
| ED visits after day 7 among non-completers | 2/5 (40.0%) |

Three patients received Level 1 health advice (3 notifications, no red flag symptoms),
One patient received Level 2 health advice (1 notification, 1 red flag symptom),
\*One patient received Level 2 followed by Level 3 health advice (2 notifications, each with 1 red flag symptom)
Two patients received Level 3 health advice (5 notifications, 4 with 1 or more red flag symptoms).

Of the 130 combined questionnaires sent (130 symptom, 75 PRO), 78 symptom and 45 PRO questionnaires were completed (60% questionnaire completion rate). The post-discharge completion rates ranged from 55.0% to 66.7%: 55% on Day 1; 65% on Day 2; 60% on Day 3; 55.6% on Day 4, 66.7% on Day 5; 58.8% on Day 6, and 58.8% on Day 7. On days when both questionnaires were sent (Days 1, 3, 5, 7), both were completed in all cases. The mean (SD) symptom questionnaires completed per patient was 3.9 (3.04). The mean (SD) number of PRO questionnaires completed per patient was 2.25 (1.71). Ten (66.7%) of the 15 patients who participated after leaving the hospital completed the final (Day 7) questionnaire.

Seven patients received a total of 11 health advice notifications upon reporting new or worsening symptoms. For the 3 patients who received Level 3 health advice, a total of 3 escalation emails were sent to the attending and PCP. All 4 (57.1%) of the 7 patients who received Level 2 or 3 health advice upon reporting red flag symptoms (gastrointestinal problems, swollen arms or legs, dizziness or lightheadedness) had chart-review confirmed visits to the emergency room within the first week after discharge. None of the 13 patients who did not report any new or worsening symptoms (n=8) or submit a questionnaire (n=5) visited the emergency room during the first week after discharge, confirmed by chart review. Among the five patients who did not submit any questionnaires, two had chart-review confirmed visits to the emergency room on post-discharge day 10.

Of the 15 patients submitting one or more PRO questionnaires after discharge, physical health scores increased in 10, did not change in 3, and decreased in 2 (mean (SD) change: 6.31 (7.35)). Mental health scores increased in 8, were unchanged in 3, and decreased in 4 (mean (SD) change: 1.56 (6.66)).

Eleven patients (55%) completed follow-up interviews and offered feedback (**Table 5**). Most described the questionnaires as clear, understandable, and helpful, with no technical issues noted. Patients did not differentiate between symptom and PRO questionnaires. Some were uncertain about identifying new versus worsening symptoms, and a few occasionally missed questionnaires due to fatigue or scheduling conflicts. Clinicians identified a need for clearer instructions in escalation notifications, including clarification of roles for the discharging attending and PCP, and suggested that distinctions between new, worsening, and chronic symptoms and critical information should be emphasized. Lastly, clinicians reported confusion in interpreting PRO score trends in general and during care transitions.

**Table 5.**
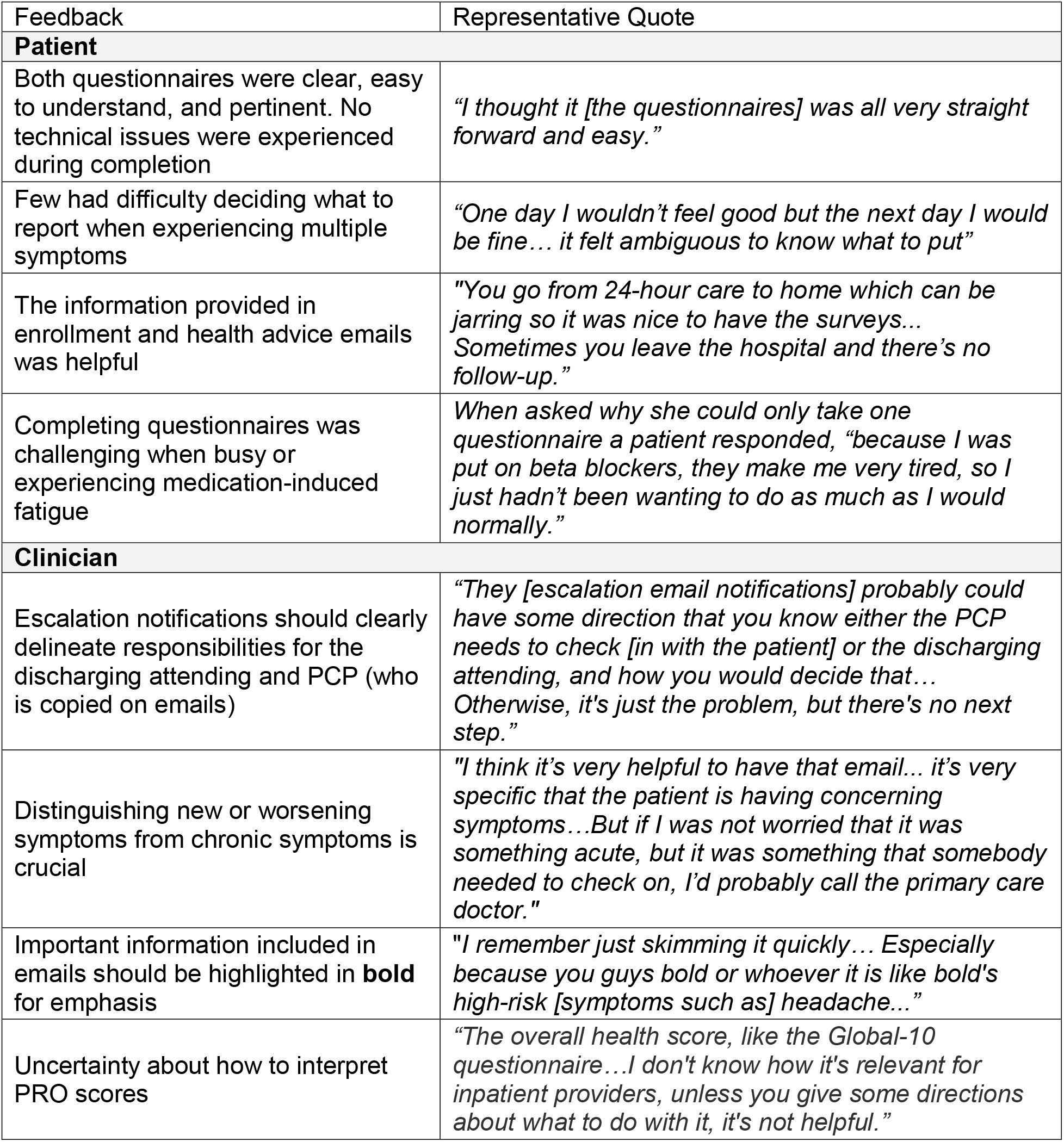
Patient and Clinician Feedback from Field Testing.

## DISCUSSION

In this mixed methods study, we used an iterative user-centered approach to develop and field test MyPostDischargePal, an automated remote monitoring intervention for adults with MCCs during the transition home following hospitalization. The intervention incorporated combined symptom monitoring, serial PROMIS Global-10 assessments, personalized risk-stratified health advice, and clinician escalation for high-risk symptoms. Patient and clinician feedback informed refinement of key features, including clearer communication of health risks, actionable guidance aligned with discharge instructions, and a ‘smart’ notification system to workflow-sensitive escalation pathways. Field testing demonstrated the feasibility and acceptability of collecting patient-reported symptoms and PRO data during the first week after discharge. Although patients found the questionnaires clear and useful, clinicians identified interpretation of longitudinal PRO trends as an important area requiring additional support and refinement.

A central finding was that patients wanted clear communication regarding post-discharge risks accompanied by actionable next steps. In response, the intervention incorporated explicit definitions of new or worsening symptoms, identified appropriate clinical contacts—the discharging attending for hospitalization-related concerns and the PCP for ongoing management—and delivered personalized health advice aligned with discharge instructions. Because approaches to communicating risk during high risk care transitions remain underexplored,[41 42] ‘health advice’ was intentionally framed using a familiar color-coded schema (green, yellow, red) corresponding to increasing urgency. In our feasibility sample, 4 of the 7 participants who received Level 2 or 3 health advice visited the ED within one week of discharge. This observation should be interpreted cautiously given the small sample, absence of a comparator, and potential cofounding by illness severity. Two of the 5 patients who did not submit questionnaires had ED visits on post-discharge day 10, highlighting the need the importance of understanding non-response patterns in future evaluations.

From the clinician perspective, clear delineation of responsibilities between inpatient and outpatient physicians emerged as a key requirement, consistent with prior literature on care transitions.[39 43 44] To support continuity, escalation notifications communicated relevant discharge information, anticipated issues, and patient-reported concerns while clarifying expected roles for discharging attending and PCPs.[38–40] Clinicians nevertheless expressed concerns regarding alert fatigue and workflow burden. To balance patient safety with acceptability, escalation notifications were limited to the first occurrence of a Level 3 event and copied to a centralized study inbox for monitoring.[45] Future implementations—including those that leverage artificial intelligence—should evaluate direct integration into EHR messaging systems and escalation pathways that more closely align with existing clinical workflows.[46–48]

Our findings align with emerging literature suggesting that effective remote monitoring interventions target high-risk populations, support self-management, facilitate timely recognition of health declines, and integrate patient-generated data into clinical care.[49] Importantly, this work extends remote monitoring to adults with MCCs discharged home from general medical services, a population at particularly high risk for post-discharge AEs.[50] Patient feedback suggested that participants valued structured follow-up during the vulnerable transition from hospital to home—“You go from 24-hour care to home which can be jarring so it was nice to have the surveys… Sometimes you leave the hospital and there’s no follow-up”. These findings reinforce conceptual models of patient empowerment that link support, self-management, and resilience.[51–53]

In addition to assessing new or worsening symptoms, MyPostDischargePal incorporated serial PROMIS Global-10 assessments. Although physical health scores generally improved over the seven-day monitoring period, this pilot was not designed to determine whether observed score changes represented clinically meaningful differences. Instead, these findings demonstrate the feasibility of PRO collection during the post-discharge period and suggest that longitudinal PRO data may complement symptom-based monitoring. At the same time, clinician uncertainty regarding interpretation of PROMIS score trends highlights the need for future work examining how clinically meaningful changes in PROs can be incorporated into monitoring and escalation workflows.

This study has several limitations. First, findings from field-testing were observational and hypothesis-generating rather than causal. Although the escalation algorithm was informed by prior institutional work, published literature, and expert opinion,[12 14] neither the symptom weighting scheme nor the escalation thresholds were prospectively validated. The forthcoming RCT will formally evaluate the predictive performance and clinical utility of the symptom-based escalation approach. Second, the study was conducted at a single academic medical center using an English-language intervention and enrolled a predominantly White population, which may limit generalizability. Future iterations should evaluate multilingual approaches, low-literacy adaptations, caregiver-assisted or proxy participation, SMS- or telephone-based outreach, and alternative communication modalities to reduce barriers related to digital access and engagement. Third, hospital attendings—often listed as the ‘person to contact’—expressed reluctance to remain the primary contact beyond 72 hours after discharge, underscoring persistent workflow challenges common to care transitions.[19 27 29 34] While many acknowledged responding to queries after discharge and lack of institutional policy, we chose not to require physician callbacks to promote acceptance of the new transitions workflow. Fourth, although serial PROMIS Global-10 assessments were successfully collected, this pilot was not designed or powered to determine whether observed score changes represented clinically meaningful differences. A formal mixed methods evaluation of patient and clinician perspectives on physical and mental health scores and trends will be conducted during the main trial.[54]

In summary, we identified and evaluated core requirements for a remote monitoring intervention designed to support post-discharge surveillance among adults with multiple chronic conditions. The resulting prototype integrated symptom monitoring, patient-reported outcomes, personalized health advice, and clinician escalation within a workflow informed by patient and clinician needs. Findings from this feasibility study support further evaluation in a randomized controlled trial designed to assess predictive performance and clinical utility of symptom-based escalation for post-discharge adverse event surveillance in this high risk population.

## Data Availability

All data produced in the present work are contained in the manuscript

## PROTECTION OF HUMAN SUBJECTS & ETHICS APROVAL

The study was reviewed and performed in compliance with the Mass General Brigham Institutional Review Board (2021P002593)

## Supplementary Materials

**Supplementary Materials 1.**
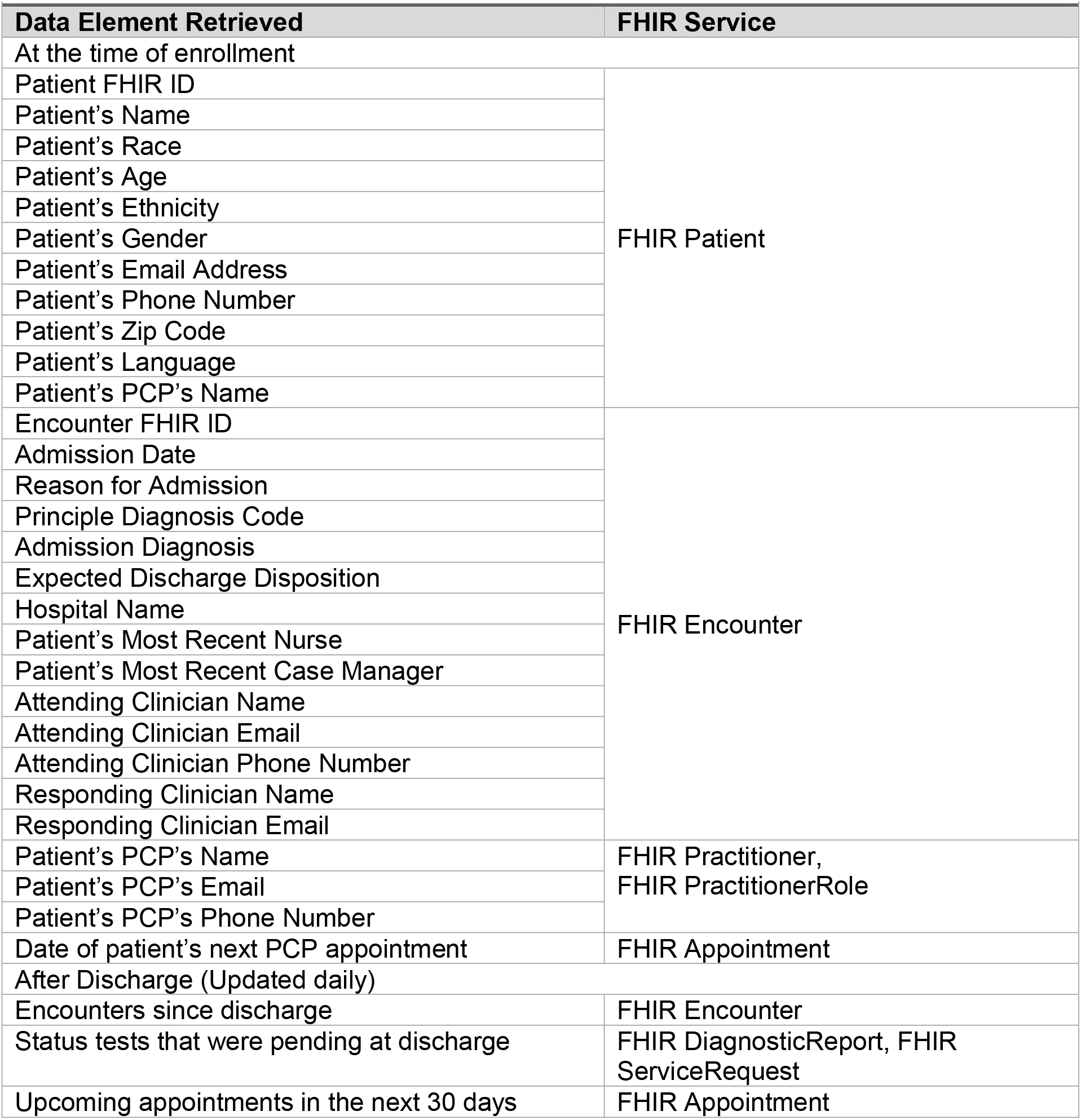
Interoperable data element & FHIR (R4) service.

